# Perceptions of the Adult US Population regarding the Novel Coronavirus Outbreak

**DOI:** 10.1101/2020.02.26.20028308

**Authors:** SarahAnn M. McFadden, Amyn A. Malik, Obianuju G. Aguolu, Kathryn S. Willebrand, Saad B. Omer

**Affiliations:** Yale Institute for Global Health, New Haven, CT 06510, USA; Yale School of Medicine, New Haven, CT 06510, USA; Yale School of Public Health, New Haven, CT 06510, USA

## Abstract

**Background:** COVID-19 outbreak is spreading globally. Although the risk of infection in the US is currently low, it is important to understand the public perception of risk and trust in sources of information to better inform public health messaging. In this study, we surveyed the adult US population to understand their risk perceptions about the COVID-19 outbreak.

**Methods and Findings:** We used an online platform to survey 718 adults in the US in early February 2020 using a questionnaire that we developed. Our sample was fairly similar to the general adult US population in terms of age, gender, race, ethnicity and education. We found that 69% of the respondents wanted the scientific/public health leadership (either the CDC Director or NIH Director) to lead the US response to COVID-19 outbreak as compared to 14% who wanted the political leadership (either the president or the Congress) to lead the response. Risk perception was low (median score of 5 out of 10) with the respondents trusting health professionals and health officials for information on COVID-19. Majority of the respondents were in favor of strict infection prevention policies to control the outbreak.

**Conclusion:** Given our results, the public health/scientific leadership should be at the forefront of the COVID-19 response to promote trust.

## Background

The current novel coronavirus outbreak, COVID-19, has spread to 28 countries and territories with tens of thousands infected and thousands of deaths (1). Although risk of contracting the disease in the United States (US) is currently low (2), it is important to understand risk perceptions about COVID-19 and trust in political and public health/scientific leadership among the US population to better inform messaging and policies (3).

## Objective

In the first study of its kind on COVID-19, our objective was to survey the adult US population to better understand their risk perceptions about the COVID-19 outbreak.

## Methods

Data were collected using an electronic questionnaire via Qualtrics® (Qualtrics, Provo, UT). Participants completed the questionnaire through the CloudResearch (4) online platform in early February 2020. We asked participants to rank who they felt should lead the US response to COVID-19. Options included the president, Congress, the Director of the Centers for Disease Control and Prevention (CDC), and the Director for the National Institutes of Health (NIH; Supplementary Material). In addition, participants completed the perceived risk scale (Cronbach’s α = 0.71) which had 10 survey-items (5-point Likert Scale: 0 = strongly disagree/disagree/neutral; 1 = agree/strongly agree). We also asked about their support for restrictive infection prevention policies and the reliability of various sources of information (Supplementary Material). Yale University Institutional Review Board approved this study. Participants provided informed consent prior to data collection.

## Results

The sample consisted of 718 adults and was similar to the US population in terms of age, gender, race, ethnicity, and education (Table 1).

**Table 1.**
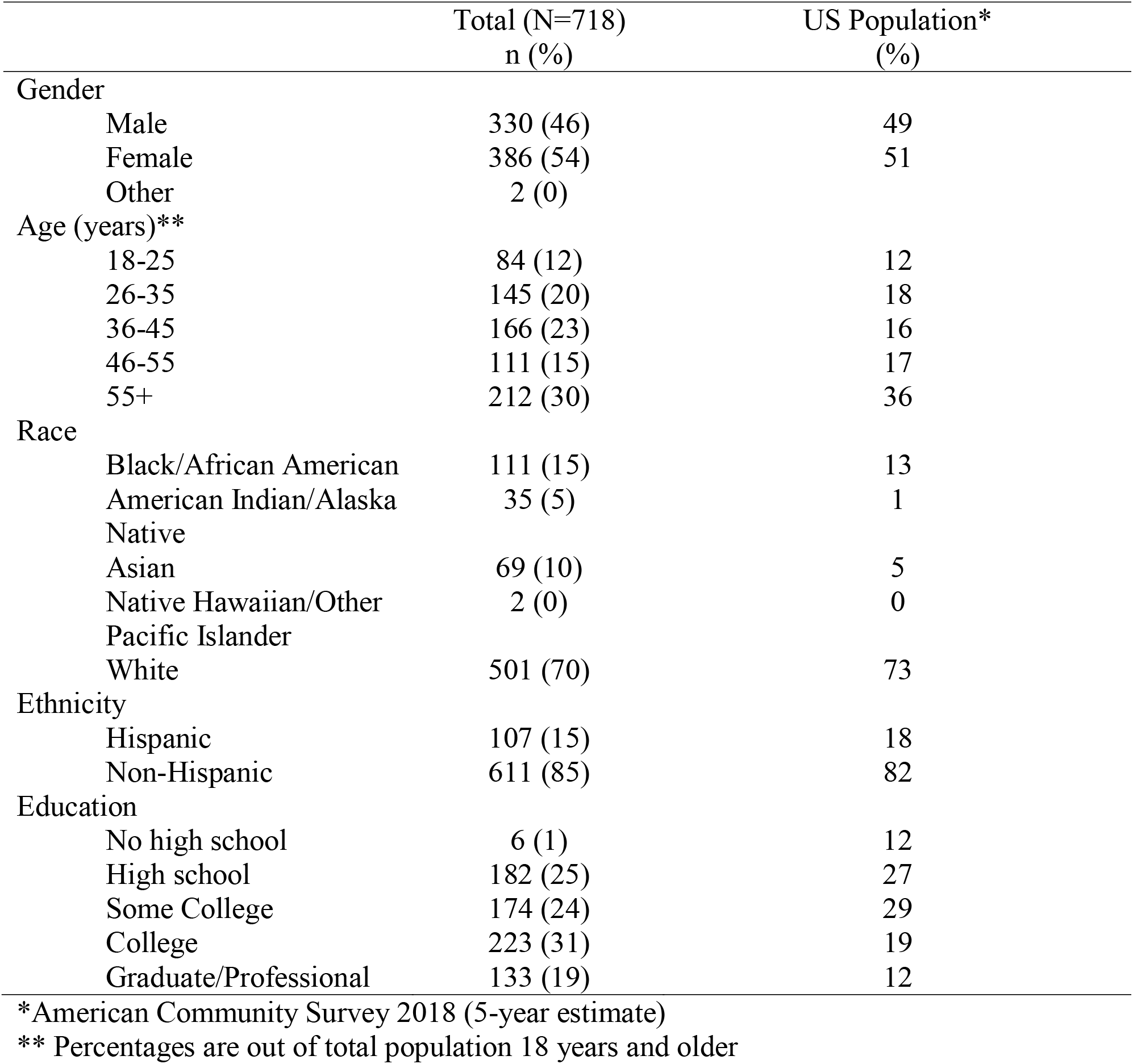
Demographic characteristics of sample compared to US population.

Over 90% of our sample was aware of the COVID-19 mostly through the news (n = 522, 73%). The majority of participants wanted the CDC Director (n = 382, 53%) or the NIH Director (n = 117, 16%) to lead the COVID-19 response (Figure 1). However, only 13% of participants wanted the president (n = 91) or Congress (n = 5, 1%) to lead the response (Figure 1).

**Figure 1.**
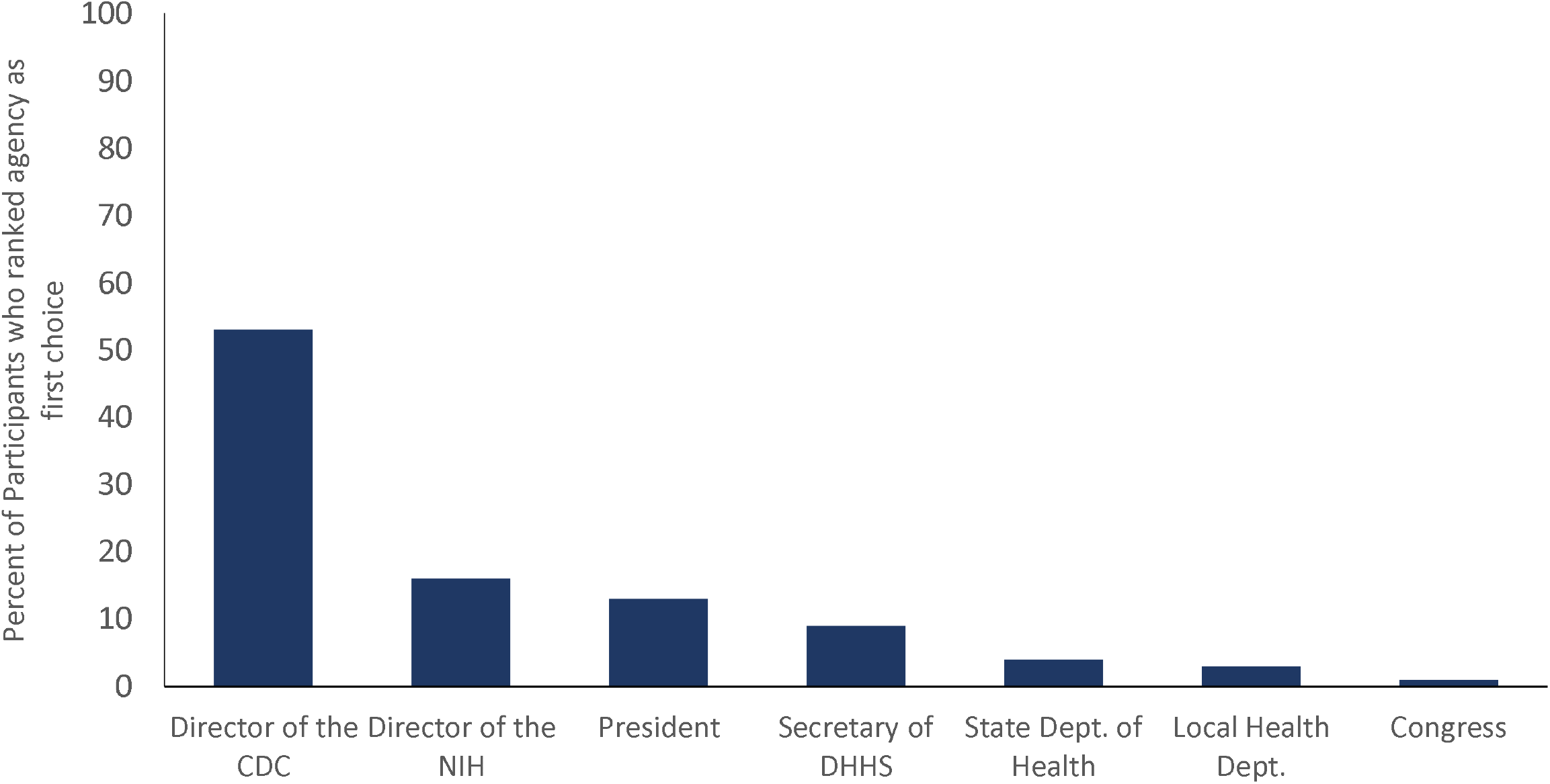
Participants choice for who should lead the US response to COVID-19 outbreak.

The mean risk perception score was 5.0 out of 10 (*SD* = 1.9; Figure 2). Strict policies for infection prevention including quarantine (n = 571, 83%) and travel restriction (n = 519, 75%) were endorsed by most participants. Additionally, thirty-five percent of participants supported “temporary discrimination based on someone’s country of origin” in case of an outbreak (n = 244, 35%).

**Figure 2.**
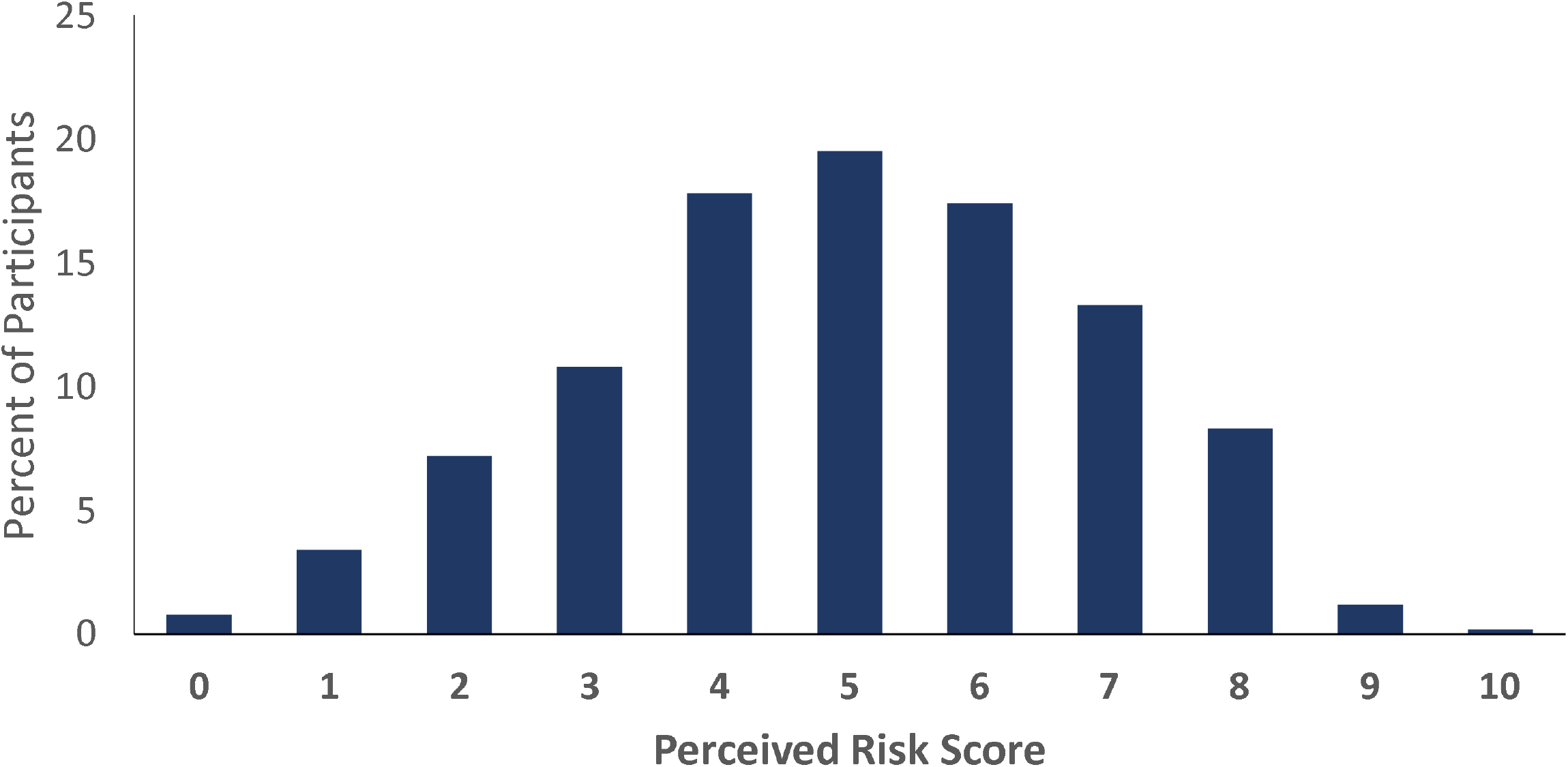
Distribution of risk perception score.

The most trusted sources of information for the participants were healthcare professionals (*M* = 4.3; *SD* = 0.9) and health officials (e.g. CDC and NIH; *M* = 4.2; *SD* = 1.0). The least trusted source of information was social media (*M* = 2.8; *SD* = 1.2; Figure 3).

**Figure 3.**
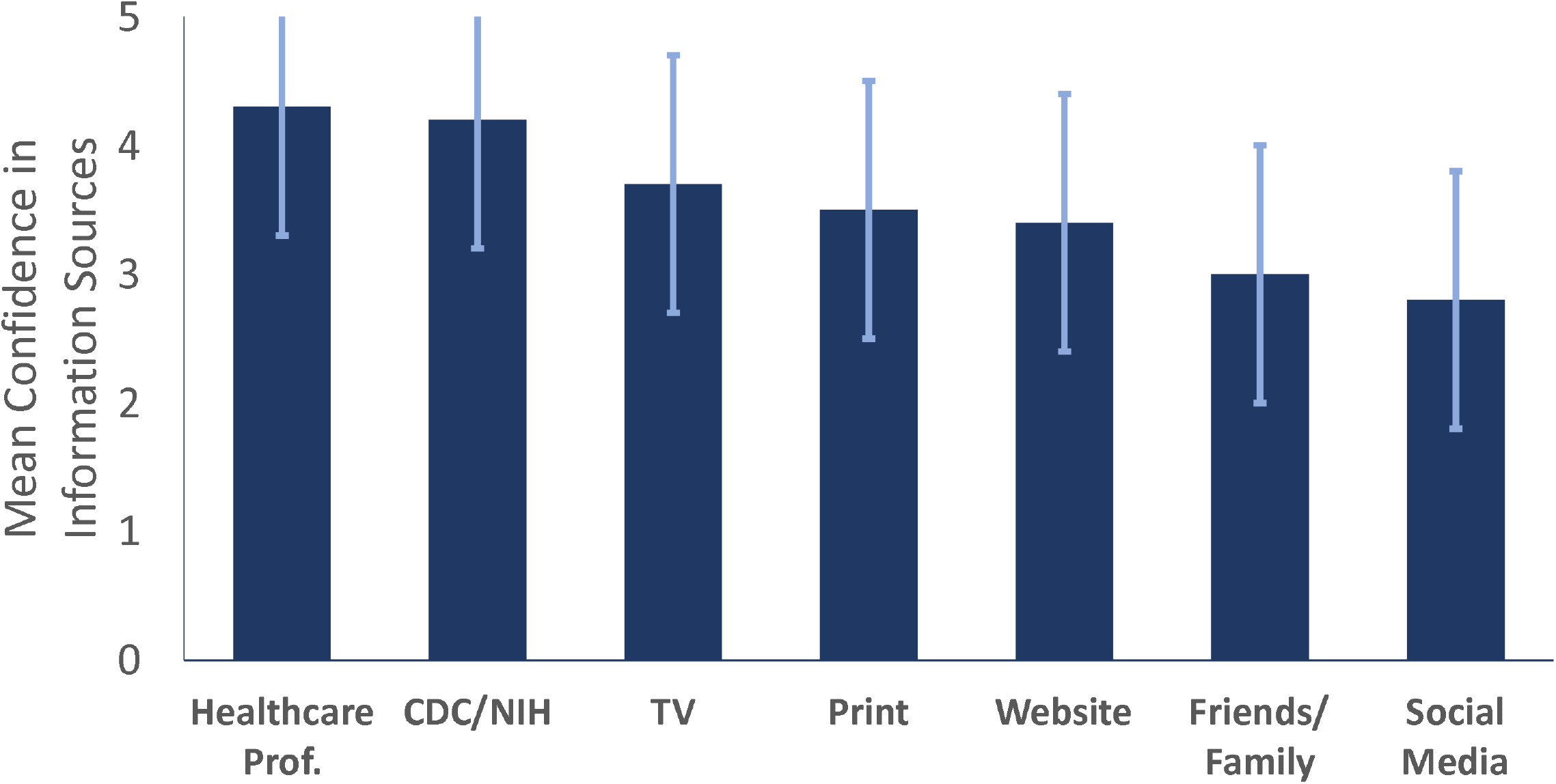
Participants confidence in various information sources.

Over 90% of the participants correctly identified CDC-recommended (5) infection prevention measures (Figure 4).

**Figure 4.**
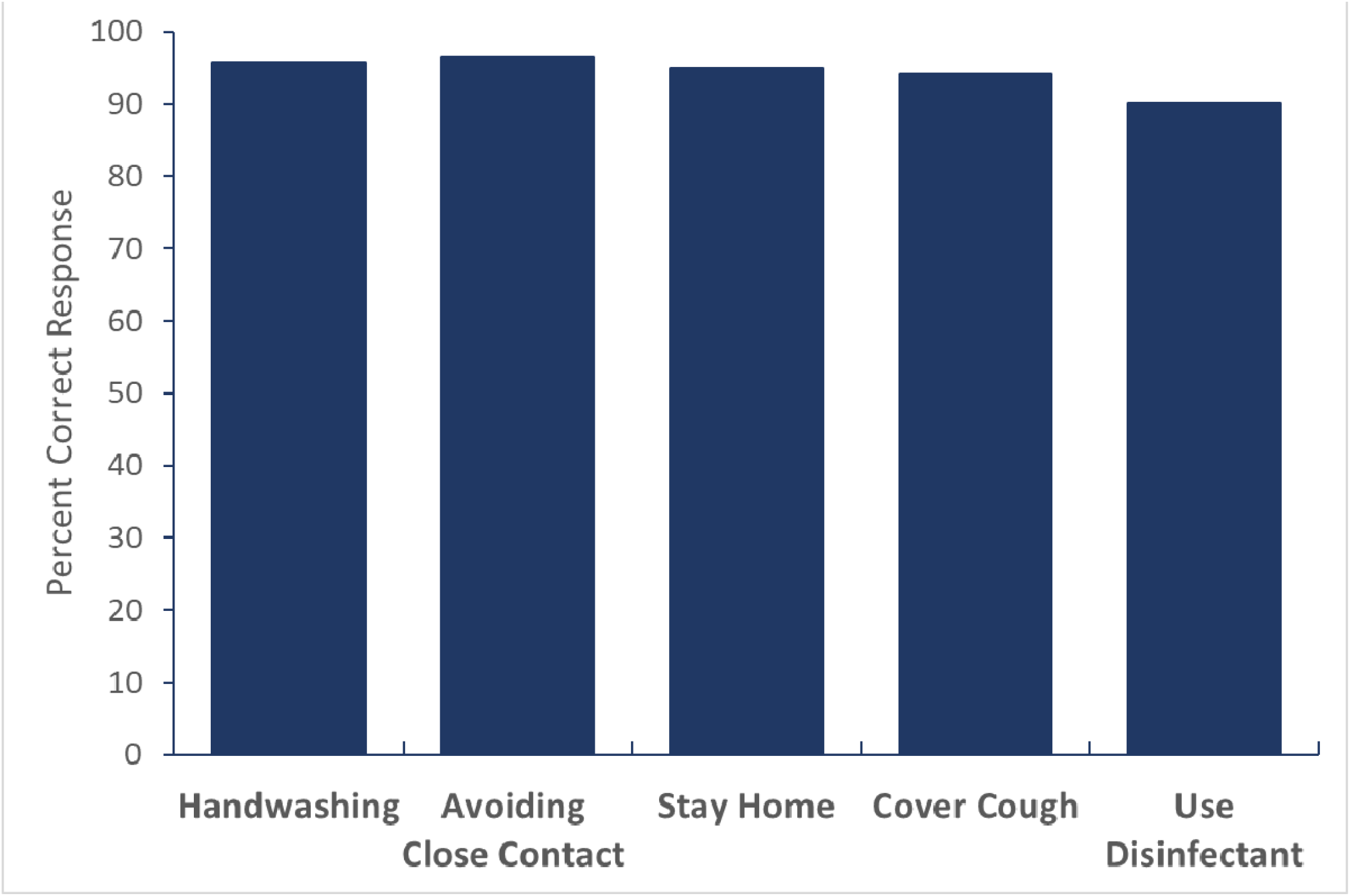
Participants Correctly Identifying Effective Infection Prevention Measures for Themselves/Others.

## Discussion

We found that the public trusted the CDC director to lead the COVID-19 response with trust in the public health/scientific leadership being high. Responsive, open, and respectful communication with the US population by these agencies may improve public health compliance and safety.^3^ Furthermore, although participants reported relatively low risk perception, many supported restrictive policies for infection prevention. A portion of the sample also supported temporary discrimination based on someone’s country of origin. These responses are concerning, and preemptive targeted messaging by the public health agencies is required to ensure a compassionate response to this outbreak. Our findings may be influenced by possible selection bias because participants needed a CloudResearch account and access to smartphone/computer to participate. However, our sample was fairly representative of the general adult US population. A weighted analysis based on age and gender demonstrate that our results are generalizable to national population (Table 2). Data for weighted analysis were extracted from US Census data.^6^

**Table 2.**
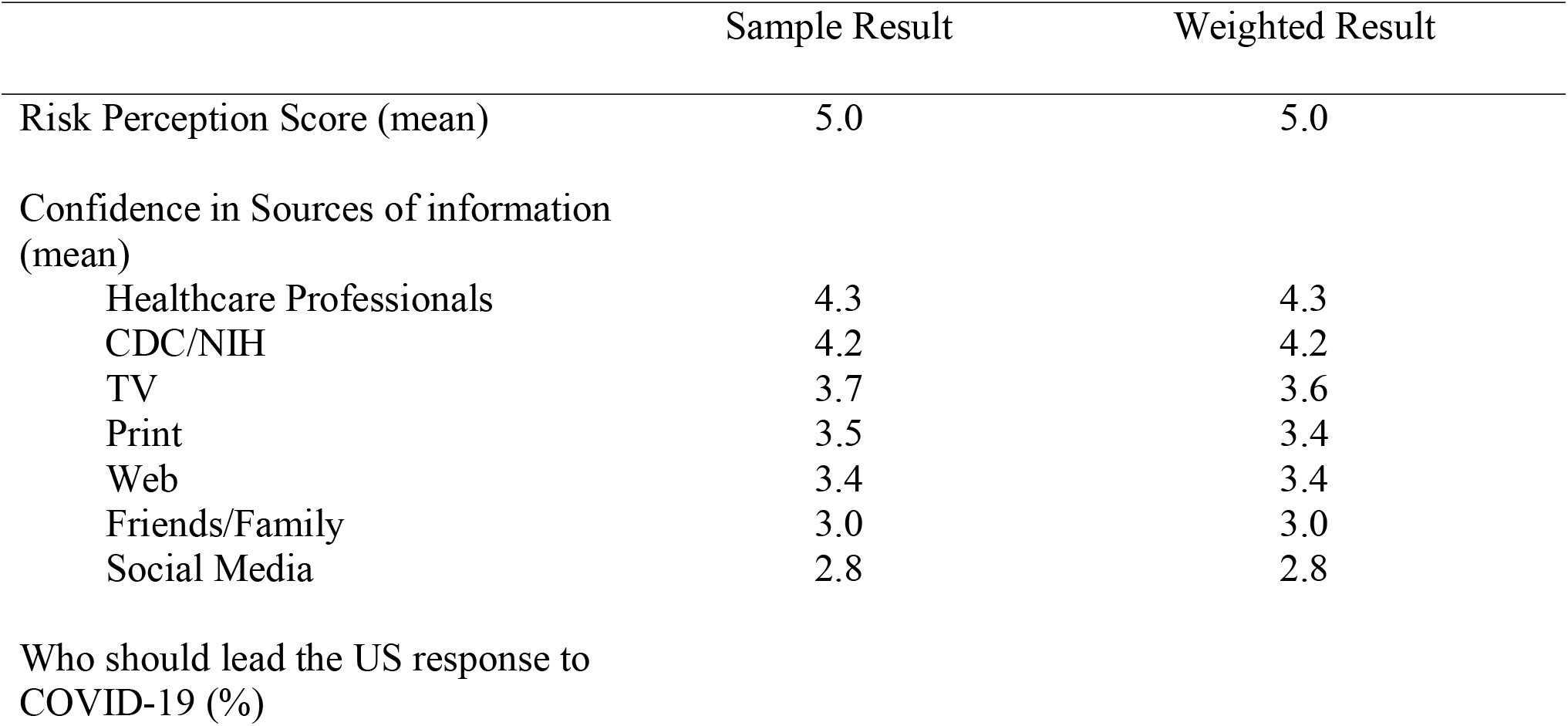
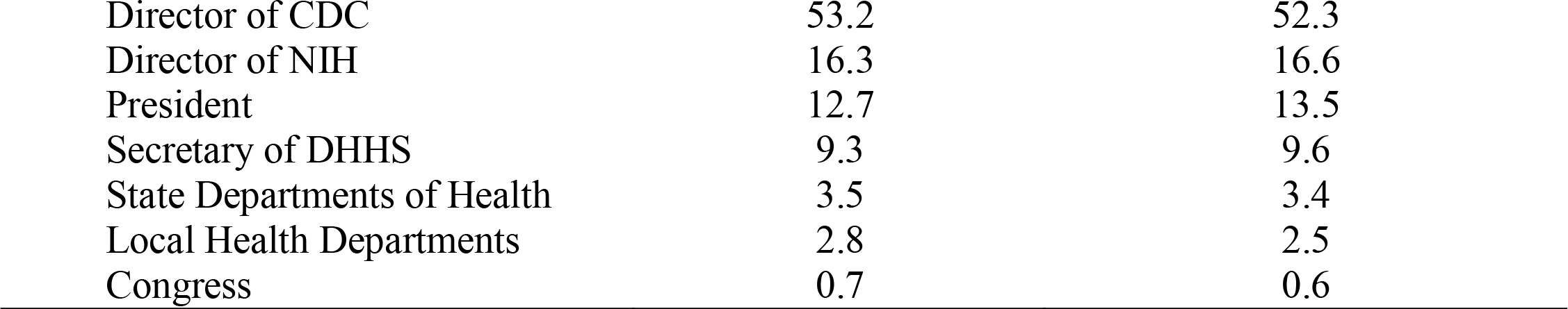
Comparison of sample result to weighted result based on age and gender.

|  | Sample Result | Weighted Result |
| --- | --- | --- |
| Risk Perception Score (mean) | 5.0 | 5.0 |
| Confidence in Sources of information (mean) |  |  |
| Healthcare Professionals | 4.3 | 4.3 |
| CDC/NIH | 4.2 | 4.2 |
| TV | 3.7 | 3.6 |
| Print | 3.5 | 3.4 |
| Web | 3.4 | 3.4 |
| Friends/Family | 3.0 | 3.0 |
| Social Media | 2.8 | 2.8 |
| Who should lead the US response to COVID-19 (%) |  |  |
| Director of CDC | 53.2 | 52.3 |
| Director of NIH | 16.3 | 16.6 |
| President | 12.7 | 13.5 |
| Secretary of DHHS | 9.3 | 9.6 |
| State Departments of Health | 3.5 | 3.4 |
| Local Health Departments | 2.8 | 2.5 |
| Congress | 0.7 | 0.6 |

Given our results, the public health/scientific leadership should be at the forefront of the COVID-19 response to promote trust. Strategic messaging by the CDC and the NIH through television, print, and internet has strong potential to alleviate unnecessary fear among the US population.

## Data Availability

Data are available upon request from the corresponding author.

## Supplemental Material

### Demographics

**Instructions: Please select the box next to the answer of your choice**.

1. **Gender**
  □ Male
  □ Female
  □ Other
2. **Age**
  □ 18 – 25
  □ 26 – 35
  □ 36 – 45
  □ 46 – 55
  □ 55+
3. **Education**
  □ No high school
  □ High school
  □ Some college
  □ College
  □ Graduate/Professional
4. **Race**
  □ Black or African American
  □ American Indian or Alaska Native
  □ Asian
  □ Native Hawaiian or Other Pacific Islander
  □ White
5. **Ethnicity**
  □ Hispanic
  □ Non-Hispanic

### Knowledge of Novel Coronavirus (COVID-19)

**Instructions: The following are questions that will ask about your knowledge level on novel coronavirus. Please select the answer of your choice**.

1. **Are you aware of the novel coronavirus outbreak?**
  □ Yes
  □ No -> *(this explanation pops up if this option is selected)* There is an outbreak of respiratory illness caused by a novel (new) coronavirus first identified in Wuhan, Hubei Province, China. There have been thousands of confirmed cases in China. Additional cases have been identified in other international locations, including the United States.
  □ Don’t know -> *(this explanation pops up if this option is selected)* There is an outbreak of respiratory illness caused by a novel (new) coronavirus first identified in Wuhan, Hubei
  Province, China. There have been thousands of confirmed cases in China. Additional cases have been identified in other international locations, including the United States.
2. **How did you learn about the coronavirus outbreak?**
  □ Media
  □ Social Media
  □ Health Officials
  □ Friends/Neighbors/Relatives
3. **How would you rate your knowledge level on novel coronavirus?**
  □ Very poor
  □ Poor
  □ Average
  □ Good
  □ Very good
4. **Which of the following is correct about the definition of novel coronavirus?**
  □ Novel coronavirus is a respiratory disease caused by a viral infection.
  □ Displayed symptoms usually include respiratory symptoms accompanied by fever, but novel coronavirus is not contagious.
  □ Novel coronavirus can progress to a severe illness but never leads to death.
  □ Don’t know
5. **Which of the following is correct about transmission route of novel coronavirus?**
  □ Novel coronavirus is transmitted through coughing or sneezing.
  □ Novel coronavirus is not transmitted by close contact with people.
  □ Don’t know.
6. **Which of the following is correct about “close contact” of novel coronavirus?**
  □ “Close contact” involves a direct contact with persons’ respiratory secretions.
  □ Relatives and healthcare workers are excluded from the category of close contact.
  □ Don’t know.
7. Which one is correct about the treatment or vaccine for the novel coronavirus?
  □ There is a curative treatment for novel coronavirus
  □ Currently, there is neither a curative treatment nor a vaccine
  □ Currently, there isn’t a curative treatment, but there is a vaccine
  □ Don’t know.
8. Which of the following are effective preventative measures for yourself and/or others against the novel coronavirus?

**8. Which of the following are effective preventative measures for yourself and/or others against the novel coronavirus?**
|  | Yes | No | Don't know |
| --- | --- | --- | --- |
| Hand washing |  |  |  |
| Avoiding touching your eyes, nose, and mouth with unwashed hands |  |  |  |
| Use of disinfectants |  |  |  |
| Staying home when you are sick |  |  |  |
| Herbal supplements |  |  |  |
| Covering your cough |  |  |  |
| A balanced diet |  |  |  |
| Avoiding close contact with someone who is sick |  |  |  |
| Use algae |  |  |  |
| Use caution when opening mail |  |  |  |
| Avoid eating meat |  |  |  |
| Getting the flu shot |  |  |  |
| Regular exercise |  |  |  |
| Wearing a face mask |  |  |  |
| None of the above |  |  |  |

### Risk Perception on Novel Coronavirus

1. **Instructions: Please select the answer depending on how much you agree with the statements below**.

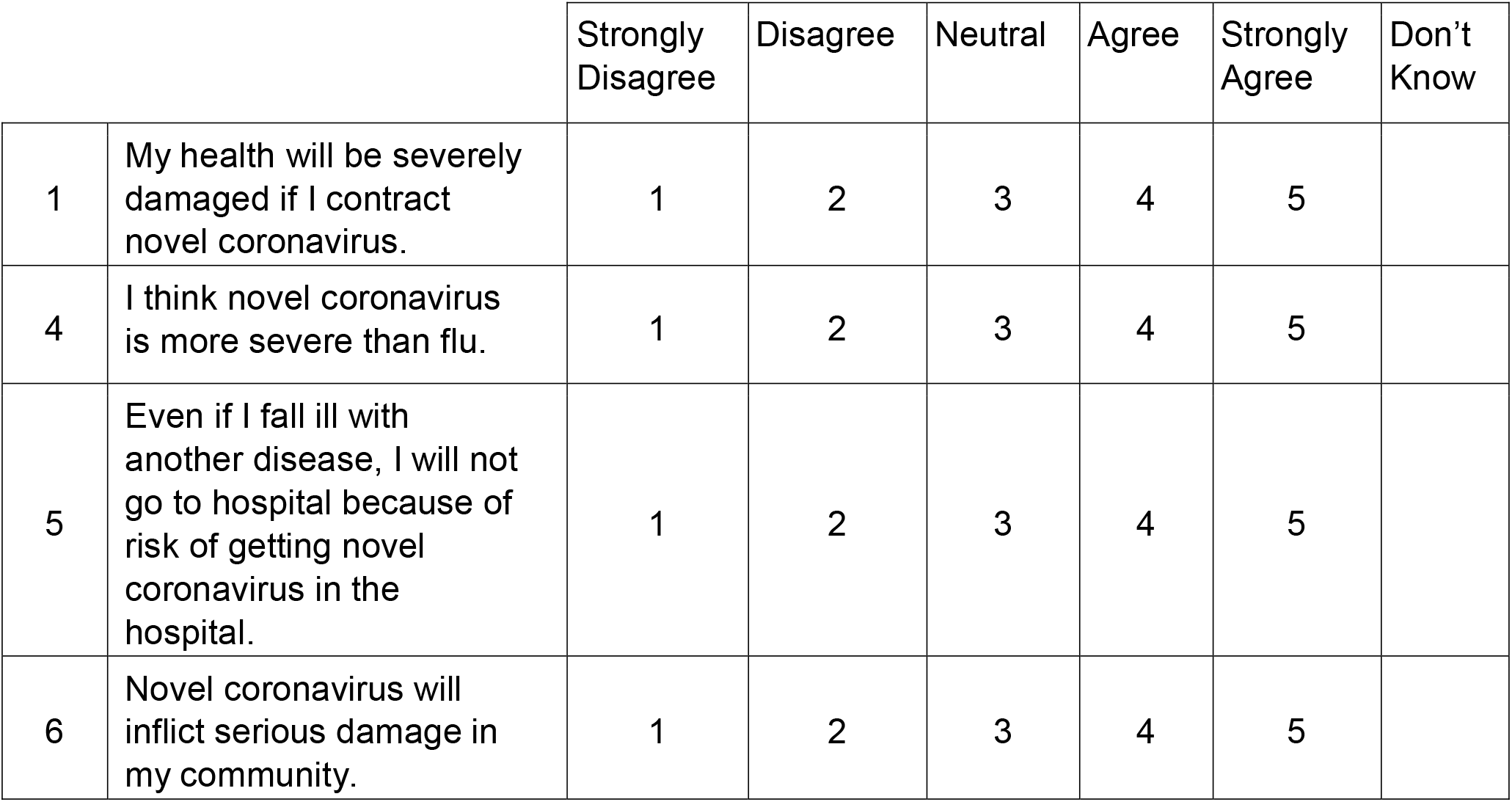
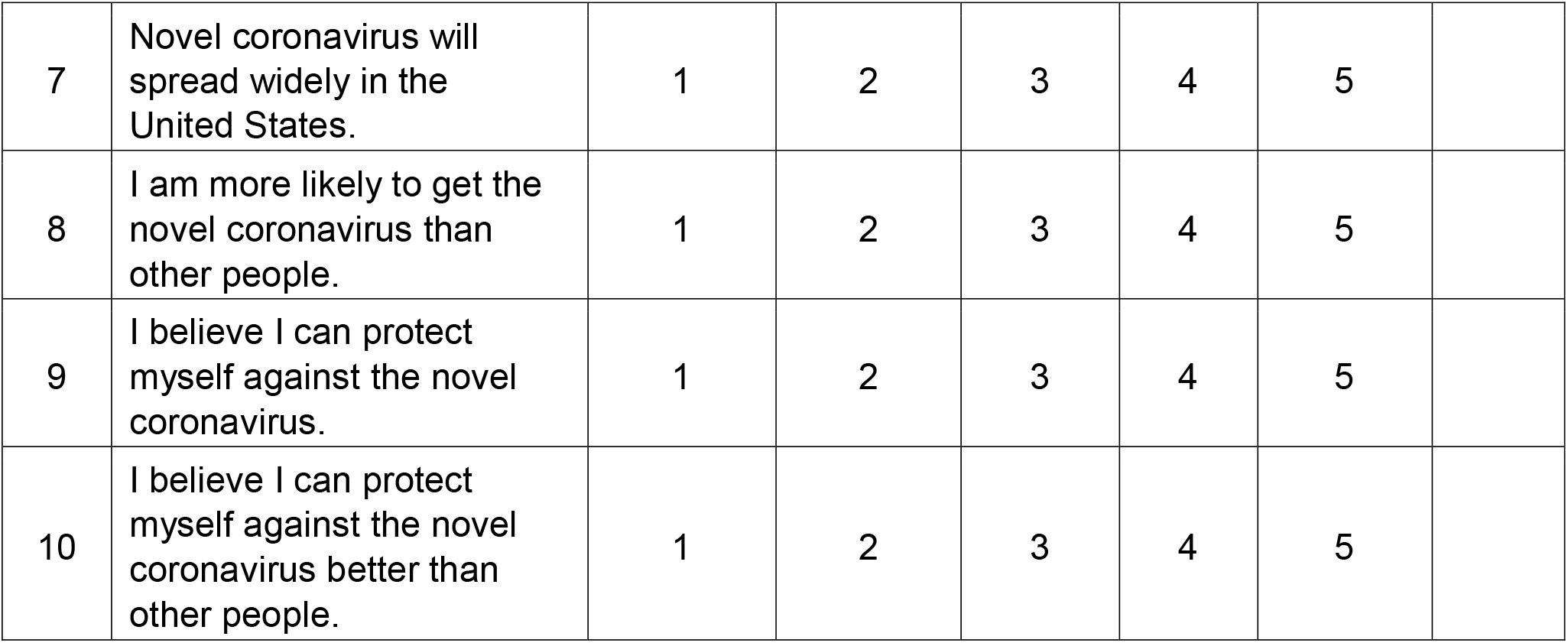
2. **What are some of the measures you have taken to prevent infection from the novel coronavirus? Select all that apply**

3. **Have you been vaccinated against the flu virus in the last 6 months?**
  □ Yes
  □ No
  □ Don’t know
4. **Do you plan to get a flu shot in the next 4 months?** *(pops up only if “no” or “don’t know” is selected for previous question)*
  □ Yes
  □ No
  □ Don’t know

**2. What are some of the measures you have taken to prevent infection from the novel coronavirus? Select all that apply**
|  | Yes | No | Don't Know |
| --- | --- | --- | --- |
| Avoided travel novel coronavirus infected areas. | Yes | No | Don't Know |
| Washed hands with soap and water. | Yes | No | Don't Know |
| Used disinfectants. | Yes | No | Don't Know |
| Avoided touching your eyes, nose, and mouth with unwashed hands. | Yes | No | Don't Know |
| Avoided eating outside of the home. | Yes | No | Don't Know |
| Stayed home when you were sick. | Yes | No | Don't Know |
| Covered your cough or sneeze with a tissue, then throw the tissue in the trash. | Yes | No | Don't Know |
| Avoided close contact with people who are sick. | Yes | No | Don't Know |
| Took an herbal supplement. | Yes | No | Don't Know |
| Exercised regularly. | Yes | No | Don't Know |
| Ate a balanced diet.<br>Used algae.<br>Used caution when opening mail.<br>Avoided eating meat.<br>Wore a face mask | Yes | No | Don't Know |
| None of the above. | Yes | No | Don't Know |

### Sources of Information

**Instructions : Please select the answer depending on how much you agree with the statements below**.

1. For the following sources of information in United States, please rate how reliable you feel they are with respect to the novel coronavirus.

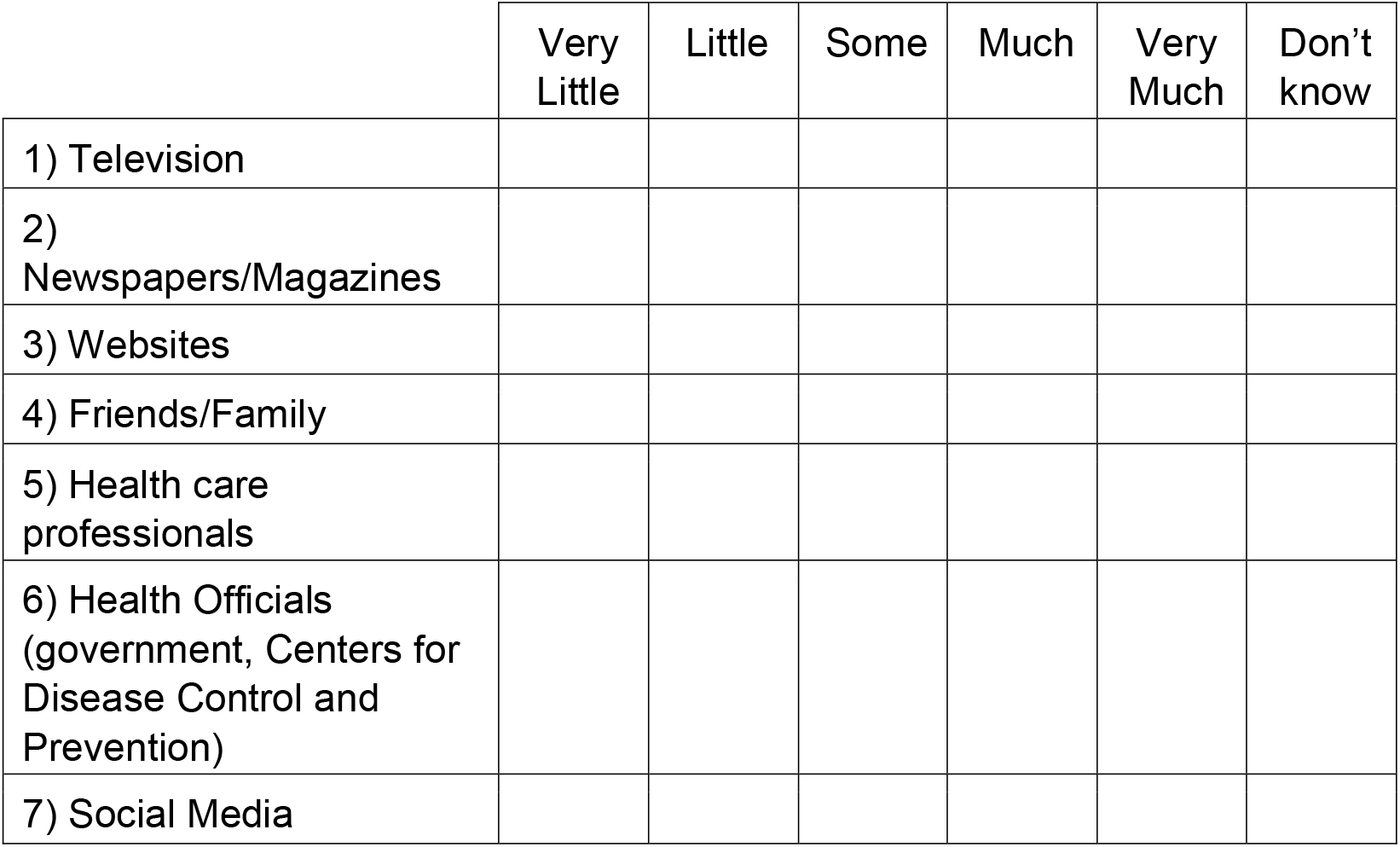

### Trust

1. How much confidence do you have in each of these organizations?

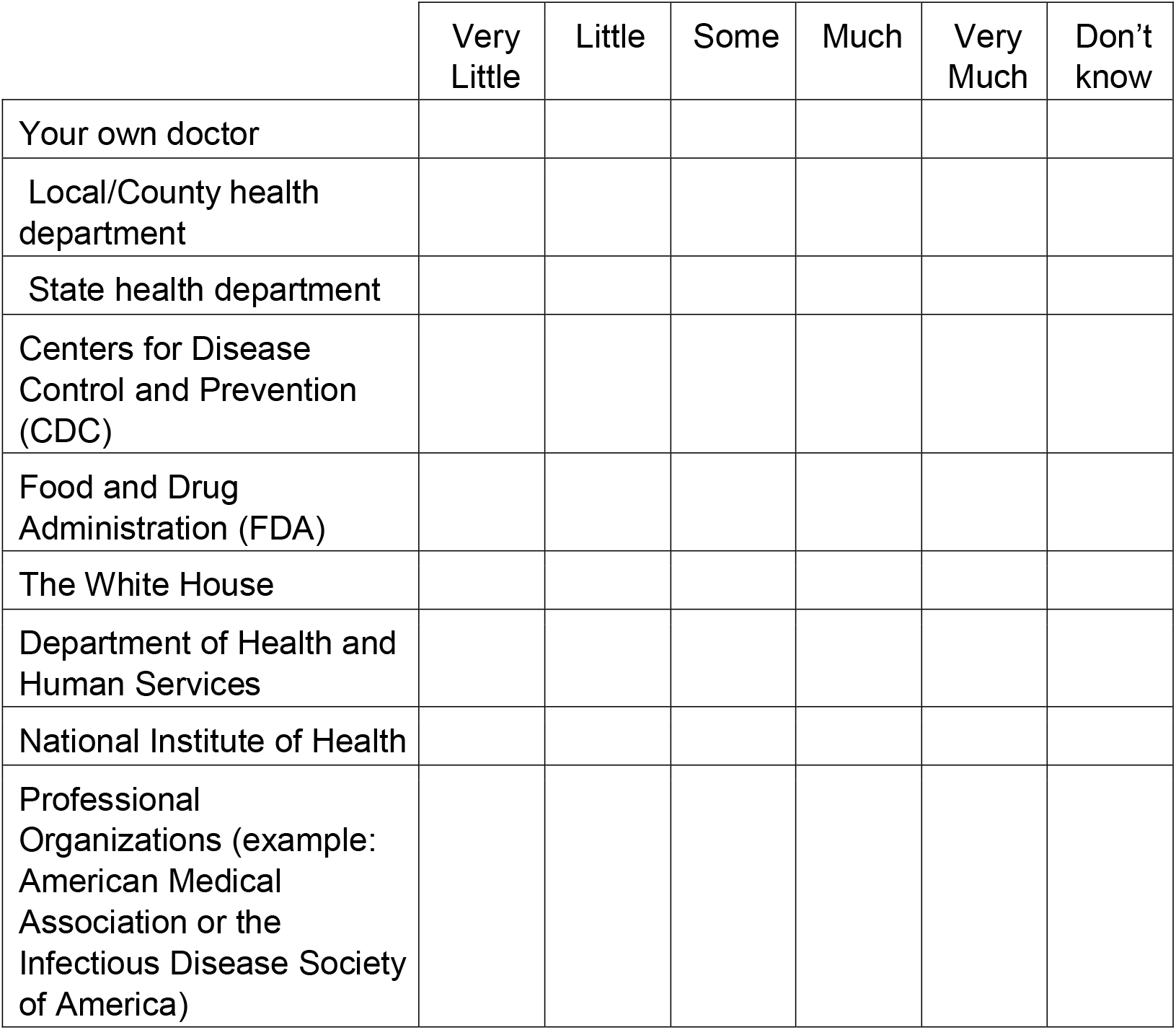
2. Please rank each of these organizations in order of your confidence in them.

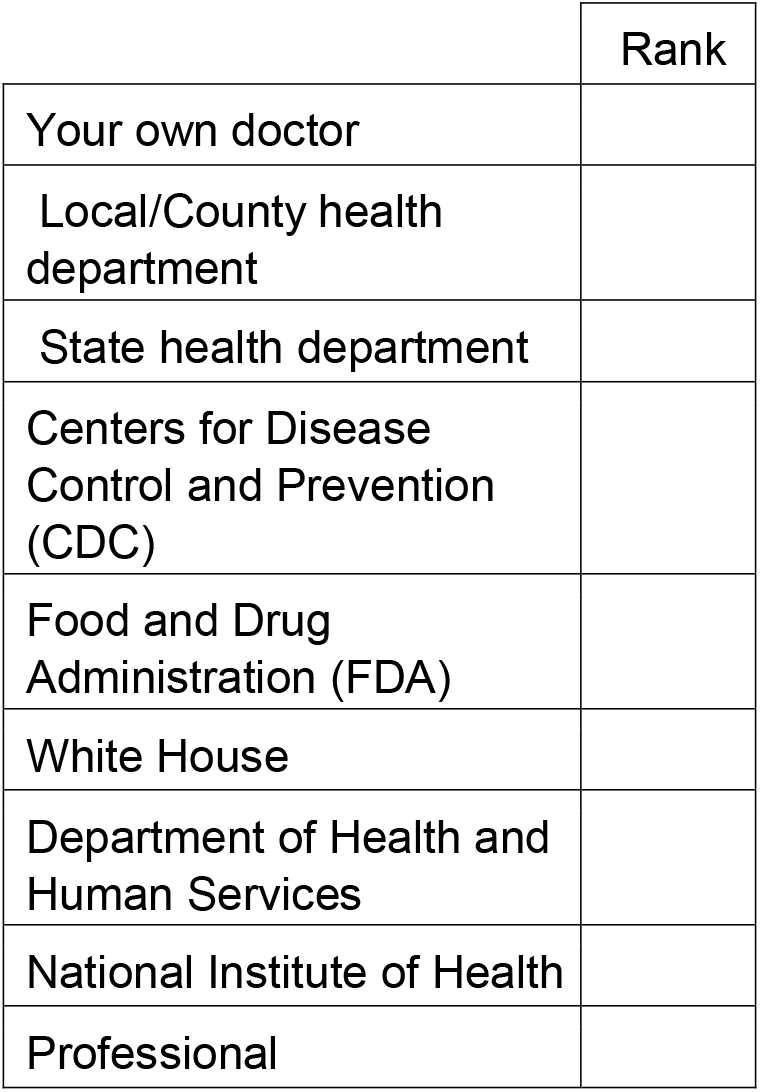
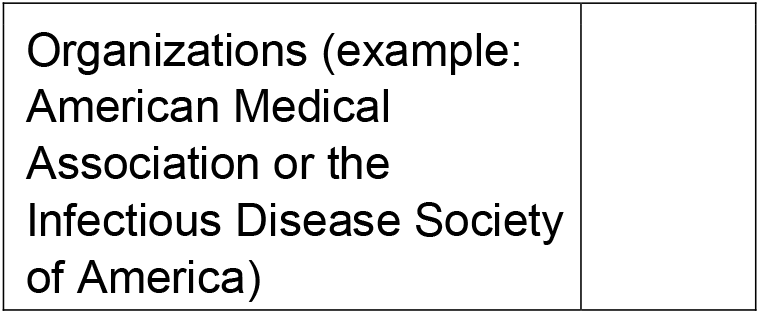

### Outrage

1. **Instructions: Please select the answer depending on how much you agree with the statements below**.

2. From your perspective, rank in order of who you think should overall be in charge of America’s outbreak response.

Outrage - 1. Instructions: Please select the answer depending on how much you agree with the statements below.**
|  | Strongly<br>Disagree | Disagree | Neutral | Agree | Strongly<br>Agree | Don't Know |
| --- | --- | --- | --- | --- | --- | --- |
| I am very concerned about this outbreak. | 1 | 2 | 3 | 4 | 5 |  |
| I believe that the government is exaggerating the threat. | 1 | 2 | 3 | 4 | 5 |  |
| I trust that health officials can effectively handle this crisis. | 1 | 2 | 3 | 4 | 5 |  |
| I trust the Congress to effectively handle this crisis. | 1 | 2 | 3 | 4 | 5 |  |
| I trust that the President can effectively handle this crisis. | 1 | 2 | 3 | 4 | 5 |  |
| I expect this outbreak to get larger. | 1 | 2 | 3 | 4 | 5 |  |
| I agree that United States should travel from China. | 1 | 2 | 3 | 4 | 5 |  |
| Americans who visited China during this outbreak should be quarantined. | 1 | 2 | 3 | 4 | 5 |  |
| In case of an outbreak, it is reasonable to temporarily discriminate against a community based on their country of origin | 1 | 2 | 3 | 4 | 5 |  |

2. From your perspective, rank in order of who you think should overall be in charge of America's outbreak response.
|  | Rank |
| --- | --- |
| The President |  |
| The Secretary of Health and Human Services |  |
| Director of National Institute of Health (NIH)/National Institute for Allergy and Infectious Diseases |  |
| Director of the Centers for Disease Control and Prevention (CDC) |  |
| State Health Departments |  |
| Local Health Departments |  |
| Other |  |
| Don't know |  |

Thank you for participating in this survey.

